# The Brazilian Rare Genomes Project: validation of whole genome sequencing for rare diseases diagnosis

**DOI:** 10.1101/2021.10.01.21264436

**Authors:** Antonio Victor Campos Coelho, Bruna Mascaro Cordeiro de Azevedo, Danielle Ribeiro Lucon, Maria Soares Nóbrega, Rodrigo de Souza Reis, Rodrigo Bertollo de Alexandre, Livia Maria Silva Moura, Gustavo Santos de Oliveira, Rafael Lucas Muniz Guedes, Marcel Pinheiro Caraciolo, Nuria Bengala Zurro, Murilo Castro Cervato, João Bosco de Oliveira Filho

## Abstract

Rare diseases affect 3.2 to 13.2 million individuals in Brazil. The Brazilian Rare Genomes Project is envisioned to further the implementation of genomic medicine into the Brazilian public healthcare system. Here we report the results of the validation of a whole genome sequencing (WGS) procedure for implementation in a clinical laboratory. In addition, we report data quality for the first 1,200 real world patients sequenced. For the validation, we sequenced a well characterized group of 76 samples, including seven gold standard genomes, using a PCR-free WGS protocol on Illumina Novaseq 6000 equipment. We compared the observed variant calls with their expected calls, observing good concordance for single nucleotide variants (SNVs; mean F-measure = 99.82%) and indels (mean F-measure = 99.57%). Copy number variants and structural variants events detection performances were as expected (F-measures 96.6% and 90.3%, respectively). Our protocol presented excellent intra- and inter-assay reproducibility, with coefficients of variation ranging between 0.03% and 0.20% and 0.02% and 0.09%, respectively. Limitations of the procedure include the inability to confidently detect variants such as uniparental disomy, balanced translocations, repeat expansion variants and low-level mosaicism. In summary, the observed performance of the test was in accordance with that seen in the best centers worldwide. The Rare Genomes Project is an important initiative to improve Brazil’s general population access to the innovative WGS technology which has the potential to reduce the time until diagnosis of rare diseases, bringing pivotal improvements for the quality of life of the affected individuals.

## Introduction

Rare diseases represent a group of over 9,000 disorders affecting an estimated 114 to 470 million patients globally (1.5% to 6.2% of the global population) ^1^. Rare diseases with genetic etiology are the leading cause of death in children and the diagnosis is challenging, but studies showed that early genetic testing leads to clear benefits by reducing the time until diagnosis, leading to better choice of therapeutic interventions, improving couples’ confidence in having children again and reducing healthcare costs ^2^.

The human genome was first mapped through the Human Genome Project (HGP), an extensive international collaboration over 13 years ^3^. Important advances in sequencing technology, such as the development of next-generation sequencing (NGS), have enabled the sequencing of a complete genome within hours, at a fraction of the initial cost, which resulted in the generation of a large amount of data, and a widespread application for diagnosis and research ^4^.

NGS for DNA sequencing encompasses several approaches: whole genome (WGS), whole exome (WES) and targeted (panel) sequencing. With WGS, it is possible to read approximately all 3 billion base pairs of human genome ^5^. The falling cost, increasing ease of application and comprehensive nature of WGS is making it the ideal tool for routine use in rare disease diagnosis.

WGS can overcome many of the technical limitations of other NGS approaches, including uneven coverage and low sensitivity for the detection of copy number, structural and expansion repeat variants ^6^. In addition, it enables the identification of noncoding and mitochondrial variants ^7^. In fact, many studies have shown that WGS has high diagnostic yield and that early molecular diagnosis improves outcomes and reduces healthcare costs ^8, 9^.

The WGS workflow can be divided in three major steps: wet laboratory sample processing, bioinformatics analyses for variant calling and annotation, and correlation of the clinical and molecular findings, resulting in a medical report. The implementation of WGS in clinical laboratories thus requires critical assay design, validation and implementation of quality control measures according to specific guidelines recommendations, to ensure adequate performance prior to use in diagnostic routine ^10, 11^.

The Brazilian Rare Genomics Project envisions to further the implementation of genomic medicine into the Brazilian national public healthcare system (SUS), complementing current policies, significantly improving the diagnostic capacity for rare disorders.

Here we report the results of the development and validation of a PCR-free WGS protocol for clinical use in the project, including wet-lab workflow and bioinformatics pipelines. In addition, we document the protocol performance in the first 1,200 samples sequenced.

## Materials and methods

### Sample selection and test scope

Our validation dataset was composed of 76 samples (Supplementary Table 1). Among them, 22 were international reference samples purchased from Coriell Life Sciences (Philadelphia, PA, USA) for benchmarking and validation, including seven reference samples from Genome in a Bottle Consortium (GiaB) ^12^. The remaining 54 are samples previously characterized by other methodologies: single nucleotide polymorphism (SNP) array, array comparative genomic hybridization (aCGH), conventional karyotyping, or fluorescence *in-situ* hybridization (FISH). We intended to detect and report single nucleotide variants – SNVs, insertion/deletions – indels, copy number variants – CNVs (large deletions and duplications, chromosomal aneuploidy), and structural variants – SVs (inversions, translocations), as well as mitochondrial SNVs. Repeat expansions and mosaicism were not included in the scope of this first phase of validation.

Samples were sequenced across three independent routines. We selected two benchmark samples to assess reproducibility: the reference sample NA24385 was replicated into one routine for intra-assay reproducibility evaluation whereas NA24694 was included in all three routines for inter-assay reproducibility evaluation. Routines were independently performed by different operators.

### Research ethics statement

This study adheres to the Declaration of Helsinki principles for research in human beings and was approved by the Hospital Israelita Albert Einstein’s Research Ethics Committee (São Paulo, Brazil. Protocol number: CAAE 29567220.4.1001.0071). All individuals provided written consent for WGS testing.

### DNA extraction, quantification, and fragmentation

DNA was extracted from whole blood samples using QIAsymphony DNA Mini Kit at QIAsymphony automated system (both Qiagen, Valencia, CA, USA). The extracted DNA was eluted into a final volume of 90 μl with an elution buffer. Genomic DNA purity was evaluated using NanoDrop 2000 (thresholds: 260/280 ratio ≈ 1.8 and 260/230 ratio between 1.8 and 2.2). DNA quantification was performed with Qubit® 4 fluorometer using the Qubit® dsDNA HS assay (both Life Technologies, Carlsbad, CA, USA).

Genomic DNA was fragmented into 350 bp inserts by Covaris ME220 ultrasonicator (Covaris, Woburn, MA, USA), with the following treatment settings – DNA input: 1 μg (final volume of 55 μl, completed with resuspension buffer); peak incident power: 50 W, duty factor: 20%, cycles per burst: 200, duration: 65 seconds, and temperature set point: 20 °C.

### Whole Genome Sequencing library preparation

The paired-ends sequencing libraries were prepared using 50 μl of the fragmented DNA solution (1 μg DNA final) as input and Illumina TruSeq® DNA PCR-Free Library Prep protocol HS (Illumina Inc., San Diego, CA, USA) for Whole Genome Sequencing reagent kit, following the manufacturers’ instructions. Briefly, the protocol steps were: (1) Cleanup of fragmented DNA, (2) Repair ends and selection of library size, (3) Removal of large DNA fragments, (4) Removal of small DNA fragments, (5) 3’-ends adenylation (6) Adapter ligation, and (7) Cleanup of not-ligated fragments.

### Library quality control

For quality control of adapter-ligated fragment sizes, libraries were diluted 1:5 with water and 2 μl were evaluated in the automated electrophoresis analysis TapeStation System, with D1000 High Screen Tape (Agilent Technologies, Santa Clara, CA, USA). High-quality (ideal) libraries displayed only one peak around 900 bp (equivalent to ≈ 470 bp fragments due to the forked structures of adapter-ligated fragments) and peak molarity ≥ 300 pM. Good-quality libraries had peak molarity between 200 and 300 pM. Libraries with peak molarity ≤ 200 pM were rejected and preparation was repeated.

### Library pooling and quantification

We optimized the protocol for the pooling of a maximum of 28 sample libraries for sequencing on each NovaSeq® 6000’s S4 flow cell. Briefly, each library was quantified with Qubit® 4 fluorometer then normalized to 7 nM in a final volume of 11 μL. Then, we pooled the 28 libraries into a final volume of 308 μL (28 × 11 = 308 μL). Next, starting with 5 μL of the pooled solution as input, we performed two dilutions in a resuspension buffer (1:10 and 1:100, reaching the final 1:1,000 concentration). Four μL of the diluted pooled solution were used for real time quantitative polymerase chain reaction (qPCR) on ABI 7500 real time platform (Thermo Fisher Scientific, Waltham, MA, USA) using KAPA Library Quantification Kit (Roche, Pleasanton, CA, USA). The qPCR was performed in triplicate. In each qPCR run, six KAPA DNA standards with defined concentrations were included to produce a standard quantification curve. With the mean cycle threshold (CT) of the diluted samples, we calculated the concentration of the pooled libraries solutions via linear regression, while correcting for the size-difference of the KAPA standards in relation to the adapter-ligated fragments (452 bp versus 470 bp). Each pooled library was then normalized to 3 nM final concentration.

The pooled libraries were then spiked with 1.9 μL of 2.5 nM PhiX Control v3 reagent (Illumina Inc., San Diego, CA, USA). The pooled libraries were then denatured with 77 μL of fresh 0.2N NaOH solution, followed by homogenization by vortex (1800 RPM for 1 minute), centrifugation at 280 × g for 1 minute and incubation at room temperature for 8 minutes. Then, 78 μL of 400 mM Tris-HCl (pH 8.0) solution was added to the libraries pool to neutralize the NaOH. Once again, the pooled libraries solution was homogenized by vortex (1800 RPM for 1 minute) and centrifuged at 280 × g for 1 minute. The full volume (466.9 μL) of the PhiX-spiked denatured library pool solution was then transferred to NovaSeq® 6000 Reagent Kit tube and proceeded to sequencing.

### Sequencing

We performed sequencing with NovaSeq® 6000 platform using S4 flow cells with 300 cycles (150 for forward reads and 150 for reverse reads). Usually, each round of sequencing was composed by 28 pooled samples as described above, using both flow cells available (total 56 samples per run). Desirable sequencing quality metrics were cluster passing filter > 70% and flow cell occupation > 70%.

### Bioinformatics pipeline and quality control metrics

The raw sequencing files (base call file, BCL format) were converted to FASTQ format and demultiplexed in a single step using Illumina’s *bcl2fastq* program ^13^. Illumina’s DRAGEN pipeline version 3.6.3 was used to perform all alignment and variant call (SNVs, indels, CNVs, SVs) steps. Quality control metrics are provided during each DRAGEN run.

Desirable alignment quality metrics were percentage of bases that meet Q30 score > 90%, 20X minimum coverage for both whole genome and autosomes, uniformity of coverage ≥ 80%, median insert size > 300 bp, percentage of mapped reads > 98%, percentage of chimeric (supplementary) reads < 5%, DNA contamination ≤ 2%, and percentage of mapped reads marked as duplicate < 10%. Some of these thresholds were adopted from recommendations published elsewhere ^10^.

The DRAGEN-generated Variant Call Format (VCF) files were validated to ensure they had the correct format, and sample- and variant-specific quality metrics were also calculated. Each sample was assessed to ensure that percent autosomal callability was > 95% as suggested elsewhere ^10^.

High-quality variants were those which passed Variant Quality Score Recalibration (VQSR) filter, had read depth (RD) ≥ 10 and genotype quality (GQ) ≥ 20 in at least 80% of the individuals in the sample, their alternative alleles were present in at least one individual with RD ≥ 10 and GQ ≥ 20, and were not located into locations with high multiallelic variation (more than four alleles, includes non-pseudoautosomal region of X and Y chromosomes).

Functional annotation of the variants was performed with a proprietary tool, Varstation (https://varsomics.com/varstation/) developed by Hospital Israelita Albert Einstein (HIAE). The VCF files were uploaded into the service, whose workflow is based on ANNOVAR ^14^. The variants are then classified according to international good practices on genetic variants analyses and guidelines from the American College of Medical Genetics (ACMG) ^15^ and the Association for Molecular Pathology (AMP) ^16^.

### Data analysis

The 76 samples were separated into two different analytical groups. The first group included seven sequencing libraries corresponding to GiaB benchmark samples (NA12878, NA24385, NA24149, NA24143, NA24631, NA24694, and NA24695). The second group included the remaining 69 samples, i.e., the remaining 15 GiaB samples and the 54 in-house characterized samples.

The first group was analysed by comparison of the VCF files generated by our Bionformatics pipeline with reference VCF files provided by GiaB (version NISTv3.3.2). Each sample had an accompanying BED file with high-confidence regions coordinates. We performed the comparison through *vcfeval* software (Real Time Genomics, Hamilton, New Zealand) ^17^. Briefly, *vcfeval* quantifies the number of true positives (the variant call is present in both the reference file and our file), false positives (the variant call is absent in the reference file but present in our file) and false negatives (the variant call is present in the reference file but absent in our file). We then calculated the precision, sensitivity (recall), and F-measures with those numbers. Additionally, we stratified each file by SNVs and indels coordinates. In this step we calculated the mean of each metric mentioned above alongside 95% confidence intervals (95% CI).

The second analytical group samples were evaluated manually to assess the performance of not only SNVs detection, but also for CNVs and SVs, by comparing the pipeline output with the in-house annotation or the GiaB annotation, depending on the sample origin. Supplementary Table 1 contains a list of the samples used, quality metrics and a summary of expected and observed variant calls.

## Results

The mean sequencing yield considering all routines was 2.84 TB of data per S4 flow cell. Mean %Q30 score was 92.60% ± 1.36%, mean genomic coverage 38.96X ± 10.37X and mean uniformity was 96.31% ± 0.25%. Mean mitochondrial coverage was 7650.97X ± 5559.1X.

Variant calls from our WGS procedure yielded very high concordance with the reference samples. For SNVs, the mean F-measure (n=7 reference GiaB samples) was 99.82% (95% CI = 99.44% -100.0%), whereas for indels of any length was 99.57% (95% CI = 99.29% - 99.85%) (Table 1, Supplementary Table 2, Supplementary Figure 1).

**Table 1.** Quality metrics. Seven Genome in a Bottle Consortium gold standard samples were whole-genome sequenced and variant call was performed with our Varstation pipeline. The variant call files were then compared with the gold standard files through *vcfeval* program, which quantified true positives, false positives, and false negatives according to variants captured by each sample BED file with coordinates of high-confidence regions. Precision, Sensitivity and F-measure were then calculated.

| Target | Metric | Mean | Standard deviation | 95% confidence interval |  |
| --- | --- | --- | --- | --- | --- |
|  |  |  |  | Lower bound | Upper bound |
| SNVs | Precision | 0.9986 | 0.0011 | 0.9965 | 1.0000 |
|  | Sensitivity | 0.9979 | 0.0029 | 0.9922 | 1.0000 |
|  | F-measure | 0.9982 | 0.0020 | 0.9944 | 1.0000 |
| Indels, overall | Precision | 0.9961 | 0.0008 | 0.9944 | 0.9977 |
|  | Sensitivity | 0.9954 | 0.0021 | 0.9912 | 0.9995 |
|  | F-measure | 0.9957 | 0.0014 | 0.9929 | 0.9985 |
| Indels, 1 to 5 bp | Precision | 0.9965 | 0.0008 | 0.9949 | 0.9981 |
|  | Sensitivity | 0.9961 | 0.0019 | 0.9923 | 0.9998 |
|  | F-measure | 0.9963 | 0.0013 | 0.9936 | 0.9989 |
| Indels, 6 to 15 bp | Precision | 0.9939 | 0.0015 | 0.9910 | 0.9968 |
|  | Sensitivity | 0.9916 | 0.0028 | 0.9861 | 0.9971 |
|  | F-measure | 0.9927 | 0.0021 | 0.9887 | 0.9968 |
| Indels, ≥ 16 bp | Precision | 0.9832 | 0.0055 | 0.9725 | 0.9939 |
|  | Sensitivity | 0.9795 | 0.0082 | 0.9634 | 0.9955 |
|  | F-measure | 0.9813 | 0.0036 | 0.9744 | 0.9883 |

Our procedure worked best for small indels with length between 1 and 5 base-pairs (bp) (mean F-measure = 99.63%, 95% CI = 99.36% - 99.89%). Six to 15 bp indels yielded mean F-measure = 99.27%, 95% CI = 98.87% - 99.68% and 16-bp or more indels yielded mean F-measure = 98.13%, 95% CI = 97.44% - 98.83% (Table 1).

Our optimized WGS protocol presented excellent intra- and inter-assay reproducibility. Regarding SNVs, the intra-assay coefficient of variation (CV) of the F-measures was 0.04%, whereas the inter-assay was 0.03%. Regarding indels, the intra-assay F-measures CV was 0.16% whereas the inter-assay CV was 0.07% (Table 2).

**Table 2.** Reproducibility. The benchmark sample NA24385 was selected for intra-assay reproducibility evaluation whereas NA24694 was included in all three independent routines for inter-assay reproducibility evaluation. Coefficients of variation (CV) of quality metrics are reported.

| Reproducibility | Samples | SNVs |  |  | Indels |  |  |
| --- | --- | --- | --- | --- | --- | --- | --- |
|  |  | Precision | Sensitivity | F-measure | Precision | Sensitivity | F-measure |
| Intra-assay | NA24385 | 0.9970 | 0.9918 | 0.9944 | 0.9969 | 0.9948 | 0.9959 |
|  | NA24385-2 | 0.9963 | 0.9914 | 0.9939 | 0.9952 | 0.9920 | 0.9936 |
|  | *Mean* | *0.9967* | *0.9916* | *0.9941* | *0.9961* | *0.9934* | *0.9947* |
|  | *SD* | *0.0005* | *0.0003* | *0.0004* | *0.0012* | *0.0020* | *0.0016* |
|  | **CV (%)** | **0.0501** | **0.0322** | **0.0411** | **0.1221** | **0.1991** | **0.1607** |
| Inter-assay | NA24694 | 0.9992 | 0.9993 | 0.9993 | 0.9968 | 0.9980 | 0.9974 |
|  | NA24694-2 | 0.9994 | 0.9993 | 0.9993 | 0.9974 | 0.9986 | 0.9980 |
|  | NA24694-3 | 0.9988 | 0.9989 | 0.9988 | 0.9963 | 0.9968 | 0.9966 |
|  | *Mean* | *0.9991* | *0.9991* | *0.9991* | *0.9968* | *0.9978* | *0.9973* |
|  | *SD* | *0.0003* | *0.0002* | *0.0003* | *0.0005* | *0.0009* | *0.0007* |
|  | **CV (%)** | **0.0312** | **0.0235** | **0.0271** | **0.0551** | **0.0909** | **0.0726** |

SD – standard deviation, CV – coefficient of variation.

The pathogenic/likely pathogenic variant profile of the 54 in-house characterized samples included: 12 SNVs (eight missense, two nonsense, one splicing acceptor and another splicing donor), 65 large deletions (lengths ranging between 538 bp – 53,247,491 bp), including 29 loss of heterozygosity (LOH) regions identified by SNP array (lengths ranging between 812,863 bp and 72,740,279 bp); 22 large duplications (ranging between 6147 bp and 95,325,642 bp), three events of trisomy (chromosomes 13, 15 or 21), one insertion, four inversions, 10 translocations, two Robertsonian translocations and a single event of uniparental disomy, totalling 120 events.

All SNVs were correctly detected by our variant call procedure (F-measure = 100.0%). The detection of the single event of uniparental disomy failed (Table 3). The CNV and SV events detection performances were overall good (F-measures 96.6% and 90.3% respectively).

**Table 3.**
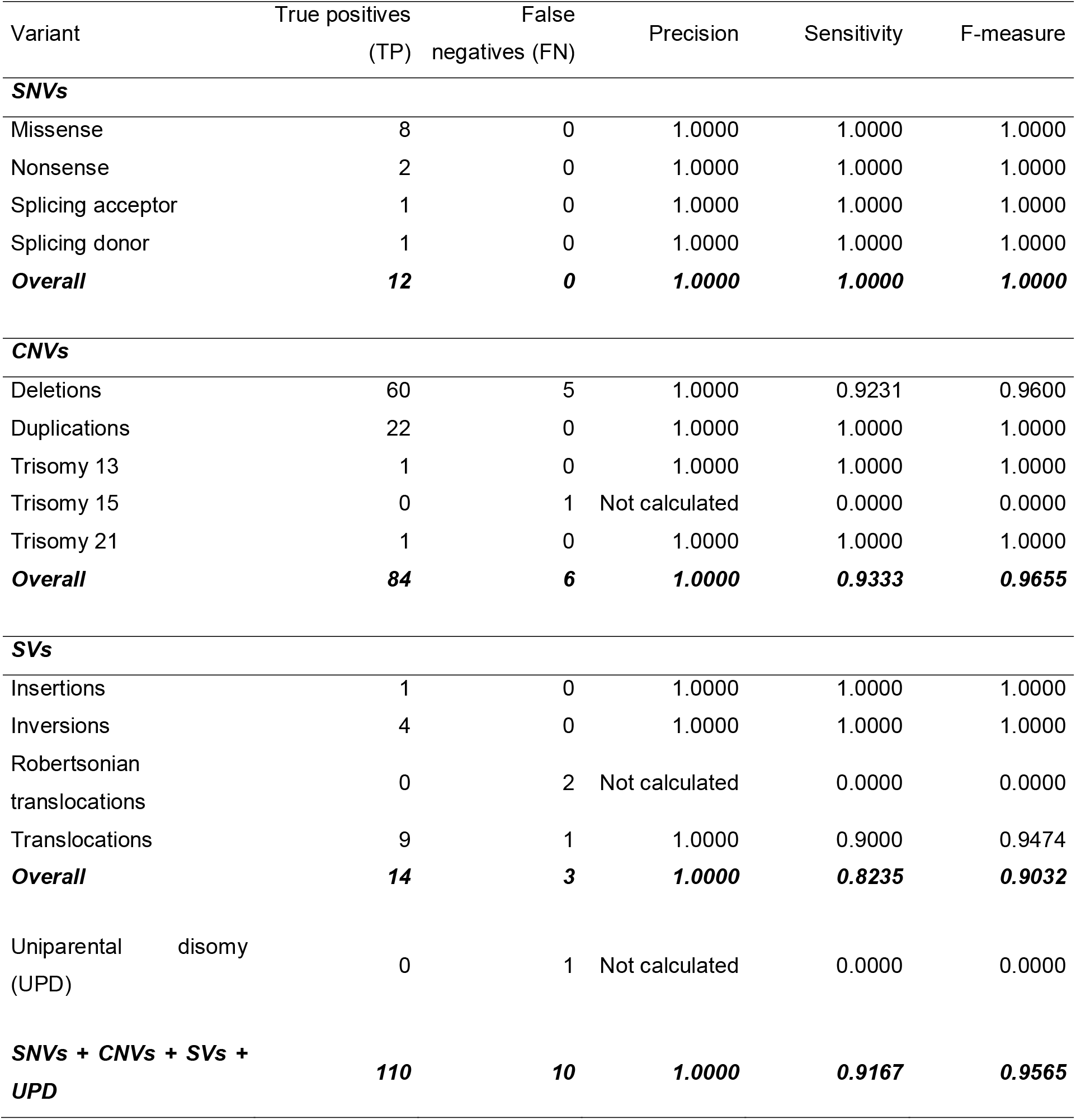
Quality metrics of the variant call procedure performed on 69 samples, including 54 in-house characterized samples by other methodologies. The seven GiaB gold-standard samples are not considered here, see Table 1. Also see Supplementary Table 1 for a breakdown of expected and observed variant calls (analysis group 2 rows).

Currently, we have sequenced over 2,000 among 3,000 enrolled patients with rare diseases or hereditary cancer syndromes with our optimized WGS protocol. Sequencing and alignment metrics are available for about 1,200 samples and have been consistently of high quality, compatible with clinical diagnostic routine (Supplementary Table 3). For example, cross-individual contamination is virtually non-existent (men 0.008% ± 0.11), the mean uniformity of coverage is 96.4% ± 0.26%, the median genome coverage is 36.5X, the mean percentage of bases with quality score Q30 or more is 91.3% ± 3.6% and mean genome callability is 96.3% ± 1.2%. Of those, over 300 patients have received a diagnostic report, with approximately 37% presenting a definitive molecular diagnosis, with the detection of a pathogenic/likely pathogenic variant compatible with the patient’s phenotype.

## Discussion

The diagnosis of patients with rare disorders is currently a lengthy process, taking on average four or more years. Early adoption of WGS could be beneficial, shortening the diagnostic odyssey ^18^. A recent meta-analysis of 37 studies involving children with genetic diseases showed that WGS testing had higher clinical utility (see^19^ for a definition) than chromosomal microarray, and an accompanying meta-regression showed that the odds of diagnosis through WGS increased by 16% each year, possibly due to methodological improvements (the meta-analysis included WGS studies published between 2015 and 2017) ^20^. Other studies reported the discovery of three new genetic conditions in children with complex phenotypes ^21^ and clinical utility for children undergoing intensive care ^22^. Therefore, WGS testing is a powerful tool with clear benefits for the diagnosis of rare diseases.

Brazil is the only country with a population larger than 100 million people which has a public, universal and free of charge health care system ^23^. Provisioning of a rapid but cost effective genomic testing strategy within a national healthcare service in order to deliver equity of access is nevertheless challenging ^24^, especially with a system of this magnitude.

As with all intensive care procedures, balancing the extreme stress of families with the complexities of an informed consent requires skill and sensitivity while incomplete understanding of a new technology creates uncertainty for care providers ^25-27^. Genomic medicine has the capacity to revolutionize the healthcare of an individual with a rare disease or cancer by offering prompt and accurate diagnosis, risk stratification based upon genotype and the capacity for personalized treatments. A recent application of WGS coupled to rare diseases diagnosis in a national context (the UK 100K Genomes Project) revealed a remarkable benefit to routine healthcare ^28^. A meta-analysis of psychological outcomes suggested no harm following WGS result disclosure and even an overall trend for a decrease in anxiety ^29^.

We developed and validated a comprehensive WGS workflow with an optimized laboratory turnaround time coupled with cutting edge Bioinformatics pipeline for variant calling, functional annotation and classification. Our procedure was performed following important benchmarking guidelines ^30, 31^ and yielded excellent performance. The key step for a robust validation is the careful sample selection. Using a set composed of reference benchmark samples, which have millions of completely validated variants of different types, and in-house characterized or purchased samples for more complex variants such as structural, mitochondrial and LOH events is of paramount importance. In addition, the validation of detection of hard-to-detect variant types, such as repeat expansions, variants in genes with pseudogenes or homologous genes and low-level mosaicism requires even further steps, including additional samples, possibly on a gene-by-gene basis ^10^.

The challenge in assessing and interpreting variants is complex, and we acknowledge some limitations of our protocol. We did not assess repeat expansion variants, tandem duplications, mitochondrial genome heteroplasmy, mosaicism, and processed pseudogene insertions. Moreover, we did not have in the sample set deletions or duplications smaller than 538 bp. Therefore, the detection sensitivity of CNVs with less than 538 bp may differ from the one reported here. We plan on validating the detection of some of these variant types in the near future, in order to improve the test robustness, sensitivity, specificity and limits of detection.

## Conclusion

Large-scale WGS projects are important initiatives to expand the population’s access to robust genomic technologies, such as NGS. The validation of our WGS workflow is the first step for this achievement and has the potential to reduce time until diagnosis of patients with rare diseases, improving the affected individuals and their family’s quality of life. Also, considering the high diversity and miscegenation of our population, The Rare Genomes Project is fundamental for the creation of a disease-related variants databases, collaborating with the future of precision medicine.

## Data Availability

The authors confirm that the data supporting the findings of this study are available within the article and its supplementary materials.

## Supplementary Figure 1 caption

Supplementary Figure 1. Circular plot displaying variant calls in the seven gold-standard GiaB samples. They are ordered according to chromosome (circular sectors) and genomic position. Each library corresponds to one circumference of said sectors. Yellow points/top third of the sector: true positive calls, red points/middle third of the sector: false positive calls, blue points/bottom third of the sector: false negative calls. Left: result for SNVs, right: result for indels.

