## Supplementary figures and images for "The Brazilian Rare Genomes Project: validation of whole genome sequencing for rare diseases diagnosis"

### Supplementary Figure 1

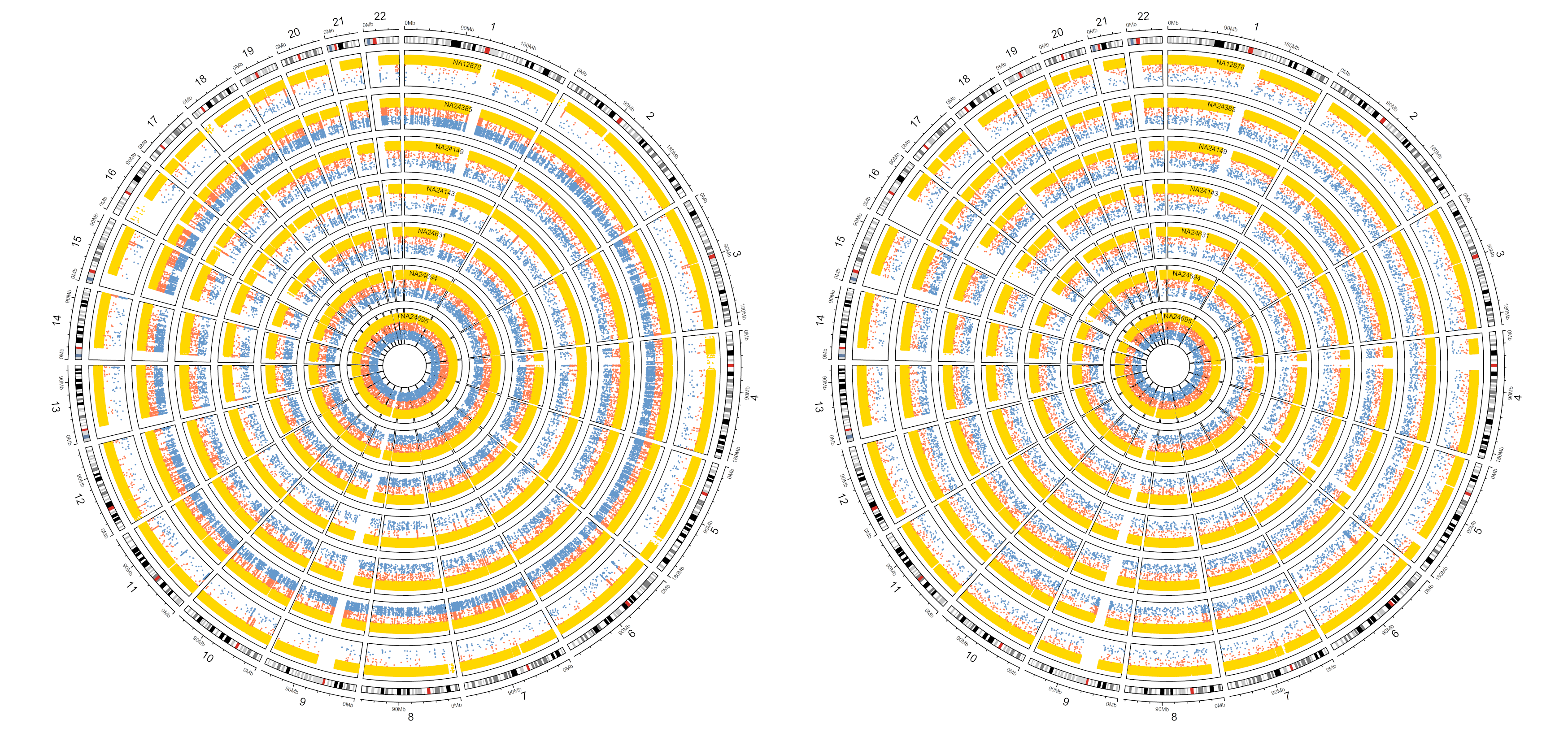
